# Interaction of inherited genetic variants in the NLRP3 inflammasome/IL-6 pathway with acquired clonal hematopoiesis to modulate mortality risk in patients with HFrEF

**DOI:** 10.1101/2023.05.03.23289436

**Authors:** Sebastian Cremer, Nikoletta Katsaouni, Wesley Tyler Abplanalp, Alexander Berkowitsch, Klara Kirschbaum, Michael A. Rieger, Steffen Rapp, Philipp S. Wild, Stefanie Dimmeler, Marcel H. Schulz, Andreas M. Zeiher

## Abstract

**Aims:** Clonal hematopoiesis (CH), defined as the presence of an expanded blood cell clone due to acquired somatic mutations in leukemia driver genes, was shown to be associated with increased mortality in patients with chronic ischemic heart failure with reduced ejection fraction (HFrEF). Mechanistically, circulating monocytes of mutation carriers display increased expression of proinflammatory genes involved in inflammasome and IL-6 signaling. Inherited genetic variants (SNP) in the IL-6 pathway are well known to affect inflammatory activation. Therefore, we investigated whether known SNPs in genes encoding for components of the inflammasome/IL-6 signaling pathway modulate fatal outcomes in HFrEF patients with CH.

**Methods and Results:** In a total of 446 patients with chronic HFrEF, peripheral blood or bone marrow mononuclear cells were analyzed for the CH driver mutations DNMT3A and TET2 as well as 48 preselected SNPs affecting genes in the NLRP3 inflammasome/IL-6 signaling pathway. The 103 patients carrying a CH driver mutation demonstrated significantly increased mortality compared to the 343 patients without CH mutations (25,24% vs 13.99% at five years; p=0.0064). We identified three commonly occurring variants known to disrupt IL-6 signaling (rs2228145, rs4129267 and rs4537545), which are in strong linkage disequilibrium and present in more than 50% of CH carriers. Harboring one of those SNPs abrogated the increased mortality risk in patients with HFrEF and CH (p≤0.05 for each SNP). On the contrary, three different SNPs (rs2250417, which is associated with increased IL-18 levels; rs4722172 and rs4845625, which are known to activate IL-6 signaling) were identified to mediate fatal outcomes in patients with HFrEF and CH; p<0.05 for each). None of the assessed SNPs influenced outcomes in patients without DNMT3A or TET2 mutations. Single Cell RNA-sequencing of circulating monocytes of patients with HFrEF revealed increased inflammatory signaling in DNMT3A mutation carriers harboring IL6/IL18 activating SNPs with genes upregulated in pathways such as “cellular response to stress”, “regulation of cell activation” and “cytokine signaling in the immune system”.

**Conclusion:** Among CH carriers with HFrEF, inherited variants in loci encoding for genes involved in inflammatory signaling interact with mortality risk. These data not only provide mechanistic insights into inflammatory mechanisms contributing to fatal outcome of HFrEF in CH carriers, but may also inform trials evaluating precision-targeted anti-inflammatory therapy in patients with DNMT3A and TET2 mutations and chronic HFrEF.

## Introduction

Clonal hematopoiesis (CH), defined as the presence of an expanded blood cell clone due to acquired somatic mutations in hematopoietic stem cells in persons without other hematologic abnormalities^1^, has recently emerged as an important independent risk factor for a variety of cardiovascular diseases.^2–4^ Specifically, CH is associated with a significantly increased mortality in patients with chronic heart failure with reduced ejection fraction (HFrEF).^5–7^ The most frequently observed CH driver mutations in patients with chronic HFrEF occur in the genes encoding for DNMT3A or TET2^8, 9^, and the association with adverse outcome appears to be independent of the presence or absence of coronary artery disease.^7^ Mechanistically, experimental studies provide robust evidence that mutations in either DNMT3A or TET2 are accompanied by increased inflammation due to increased activation of the inflammasome complex ^2, 10, 11^ and immune cell infiltration into the myocardium inducing progressive cardiac dysfunction and fibrosis.^12^ Concordant with animal and cellular models, we have previously shown by single cell RNA-sequencing techniques that circulating monocytes from patients with chronic HFrEF carrying DNMT3A or TET2 CH-driver mutations are characterized by an enhanced proinflammatory gene signature with profoundly upregulated expression of genes involved in the NLRP3 inflammasome to IL-6 signaling pathway.^13, 14^

Altered inflammatory responses are well established to contribute to the progression of chronic HFrEF^15^ and elevated IL-6 levels were shown to independently predict impaired survival in patients with chronic HFrEF.^16^ Importantly, the risk associated with enhanced inflammation with CH may be modified by inherited genetic variants within inflammatory pathways. Indeed, one recent study assessing the effects of genetically mediated reduction of IL-6 signaling in patients with coronary artery disease revealed that harboring a common variant disrupting IL-6 signaling abrogated the increased risk for the occurrence of atherosclerotic events in persons with DNMT3A or TET2 driver mutations. ^17^ However, no such studies are currently available in patients with chronic HFrEF. Therefore, we investigated the potential role of both, abrogating and enhancing genetic variants in the NLRP3 inflammasome-to-IL-6 pathway, to interfere with the prognostic significance of CH in chronic HFrEF in order to substantiate the putative involvement of inflammatory processes and provide a rationale for enhanced risk stratification for anti-inflammatory therapies.

## Methods

### Study cohort

Clinical data and biological specimens (bone marrow-derived mononuclear cells, *n* = 211, or peripheral blood cells, *n* = 235) were collected from a total of 446 patients suffering from chronic ischemic heart failure and being included in various controlled trials between January 2002 and November 2018 at the University Hospital of the Johann Wolfgang Goethe University, Frankfurt/Main, Germany.

Patients were eligible for inclusion into the study if they suffered from stable chronic heart failure symptoms NYHA ≥II, had a previous successfully revascularized myocardial infarction at least 3 months prior to inclusion, and had a well-demarcated region of left ventricular dysfunction on echocardiography. Exclusion criteria were the presence of acutely decompensated heart failure with NYHA class IV, an acute ischemic event within 3 months prior to inclusion into the registry, a history of severe chronic diseases, documented hematological disease or cancer within the preceding 5 years, or unwillingness to participate.

All patients provided written informed consent for one of the following registered clinical trials: TOPCARE-CHD (NCT00289822 or NCT00962364), Cellwave (NCT00326989), or REPEAT (NCT 01693042). Supplementary to the clinical trials, patients provided additional informed consent for genetic testing of bone marrow and/or blood samples. The ethics review board of the Goethe University in Frankfurt, Germany, approved the protocols. The study complies with the Declaration of Helsinki.

Sample preparation and next-generation sequencing was conducted as reported previously.^6^

### SNP selection, probe design, measurement, and data analysis

#### SNP selection

SNPs related to IL6 or IL1 were extracted from the GWAS catalog.^18^ Using search terms “IL6”, “IL6R” and “inflammation”, 37 SNPs were selected for the IL6 group. Using search terms “IL1A”, “IL1B” and “CASP1”, 33 SNPs were selected from related traits only (allergic disease, e.g. asthma bronchiale, hay fever or eczema, interleukin-1-beta levels, inflammatory biomarkers, protein quantitative trait loci, blood protein levels, neonatal cytokine/chemokine levels) for the IL1 group. From a total of 70 SNPs, 48 were selected for the assay according to (i) technical implementation (probe design) (ii) IL6 relation, (iii) IL1 relation, and (iv) availability on Illumina Infinium Omni EURHD genotyping chip (Illumina, Inc., San Diego, California, USA). The Infinium Omni EURHD contains approximately 2 million SNPs of the Illumina Infinium Omni 2.5-8 (Illumina, Inc., San Diego, California, USA).

#### Probe design and SNP measurements

Probes were designed by the Fluidigm Assay Design Group (Fluidigm Corporation, San Francisco, California, USA); sequences are shown in **Supplementary Table 1**. Genotyping was performed using customized SNPtype Assay, 192.24 GT Sample/Loading & SNP Kit Type™ Reagent Kit (Fluidigm Corporation, San Francisco, California, USA) according to the manufacturer’s instructions (PN 68000098 Q1, PN100-3913 C1, PN 101-8001 A1). Due to the capacity of the measuring chip (Integrated Fluidic Circuit, IFC, 24 assays/192 samples), 48 genotyping assays were splitted on two IFCs (**Supplementary Table 2)**.

Due to sufficient DNA concentration, no target amplification was carried out. For detection, 25 PCR cycles were performed on Biomark HD (Biomark Data Collection v4.7.1) using SNPType-FAM and SNPType-HEX detectors with a ROX reference. Data were postprocessed using Fluidigm SNP Genotyping Analysis v4.5.1 (Build 20180509.0910) and SNP calls were filtered by a confidence threshold of 0.9.

### scRNA-Seq Study Population and Blood Collection

Single Cell-RNA sequencing data derived from five patients with chronic HFrEF and known DNMT3A mutation status were reanalyzed with respect to SNP status. Informed consent was obtained from all patients. The study was approved by the local ethics review board and complies with the Declaration of Helsinki. Sample preparation and sc-RNA-Seq library preparation was done as reported previously.^13^

### scRNA-Seq Data Analyses

Single-cell expression data were processed using STARsolo (v 2.7.3a) to perform quality control, sample demultiplexing, barcode processing, and single-cell 3′ gene counting.

Data integration was performed by Seurat (v4.1). The gene-cell-barcode matrix of the samples was normalized by the number of unique molecular identifiers per cell. In brief, differential expression of genes in classical monocytes was used utilizing the “FindMarkers” function in the Seurat package for focused analyses. Genes with a p-value and Bonferroni adjusted p-value of *p*<0.05 were considered as differentially expressed genes. Differential transcriptional profiles by cluster were generated in Seurat with associated gene ontology terms derived from the functional annotation tool Metascape for further analyses, subset terms from Metascape analyses are shown.

### Statistical analysis

For baseline clinical characteristics, continuous variables are presented as median (interquartile range), unless otherwise noted. Mann-Whitney-U-Test was used for two group comparison of continuous variables. Categorical variables were compared with Fisher’s exact test. Error probability p< 0.05 was considered as significant. Statistical analysis was performed with SPSS (Version 24, IBM).

For survival data, statistical analysis was performed with Python (version 3.6.9), using the survival analysis library lifelines (version 0.27.0). For each of the selected SNPs the patients were divided into two groups based on the occurrence or absence of CH in the DNMT3A and TET2 genes. For the definition of CH, 1.15% and 0.73% VAF cut-offs were used for the DNMT3A and TET2, respectively.^9^ A 90% confidence threshold was set for the variant calling from the SNP array results. To avoid misconceptions, for each SNP the patients with no calls from the array were excluded for the survival analysis. After that, Kaplan-Meier analysis was conducted and log-rank testing was applied for comparison of event-free survival analysis and 95% confidence intervals (CI) were calculated. Statistical significance was assumed if *p* < 0.05.

**Table 1:**
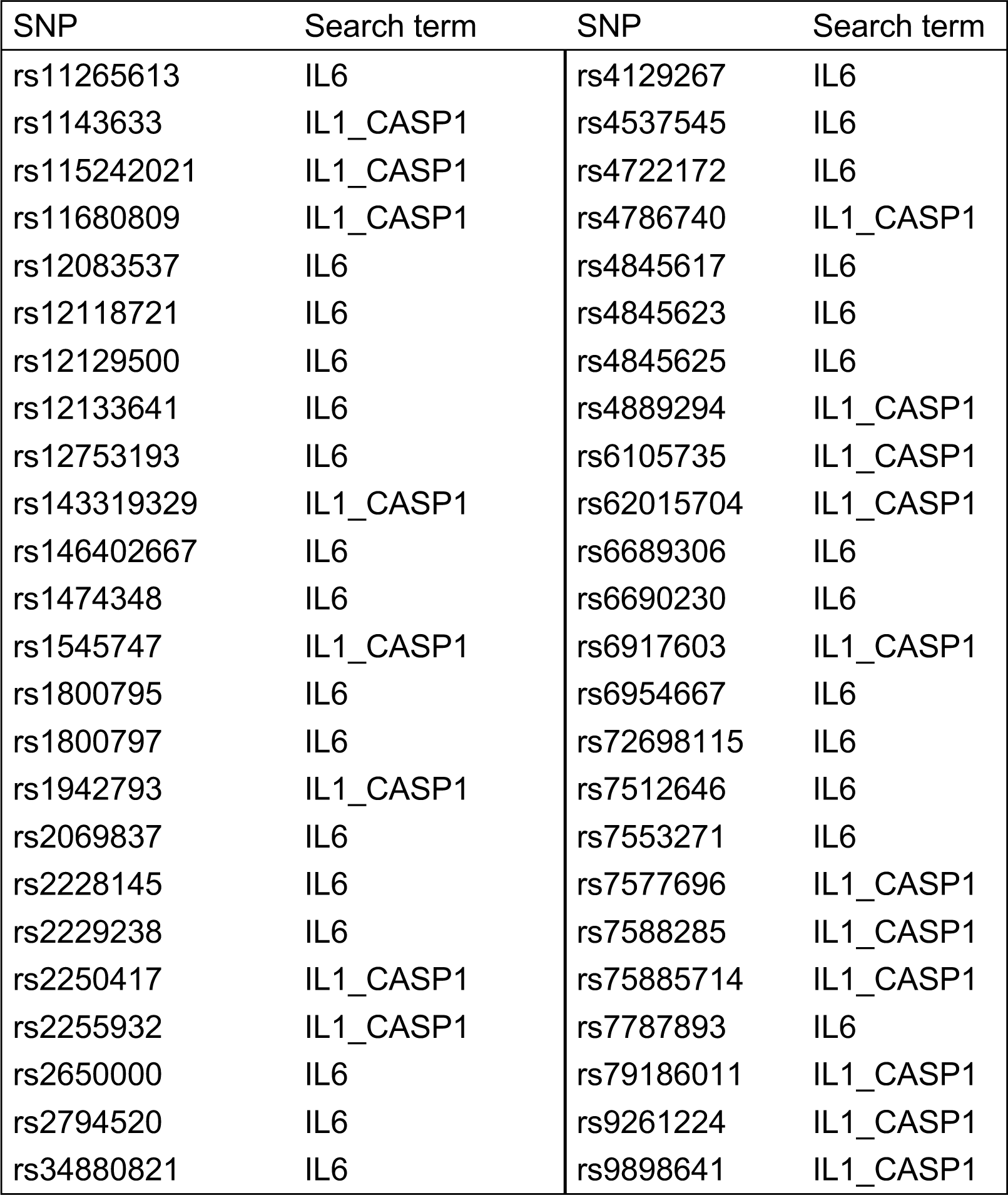
SNPs in the NLRP3 inflammasome/IL-6 signaling pathway selected for further analysis.

## Results

High sensitive error-corrected DNA-sequencing was performed in a total of 446 patients with chronic ischemic HFrEF to detect CH-driver gene mutations in either DNMT3A or TET2 in blood or bone marrow mononuclear cells as described.^6^ Median age of the patients was 63 (IQR 53-70) years. A variant allele frequency cut-off value of 1,15% for DNMT3A mutations and 0.73% for TET2 mutations was used, as we previously identified these thresholds to be associated with mortality in patients with HFrEF. ^9^ In our cohort, 103 of 446 patients had mutations above the respective thresholds. Consistent with previous reports, the presence of CH increased with age (p<0.001), was associated with higher systolic blood pressure (p=0.021) and increased mortality at five years (25,24% in CH carriers vs 13.99% in patients without mutations; p=0.0064; **Table 2, Figure 1**). There were no significant differences in other baseline parameters in patients with CH compared to patients without CH except higher use of digitalis.

**Figure 1:**
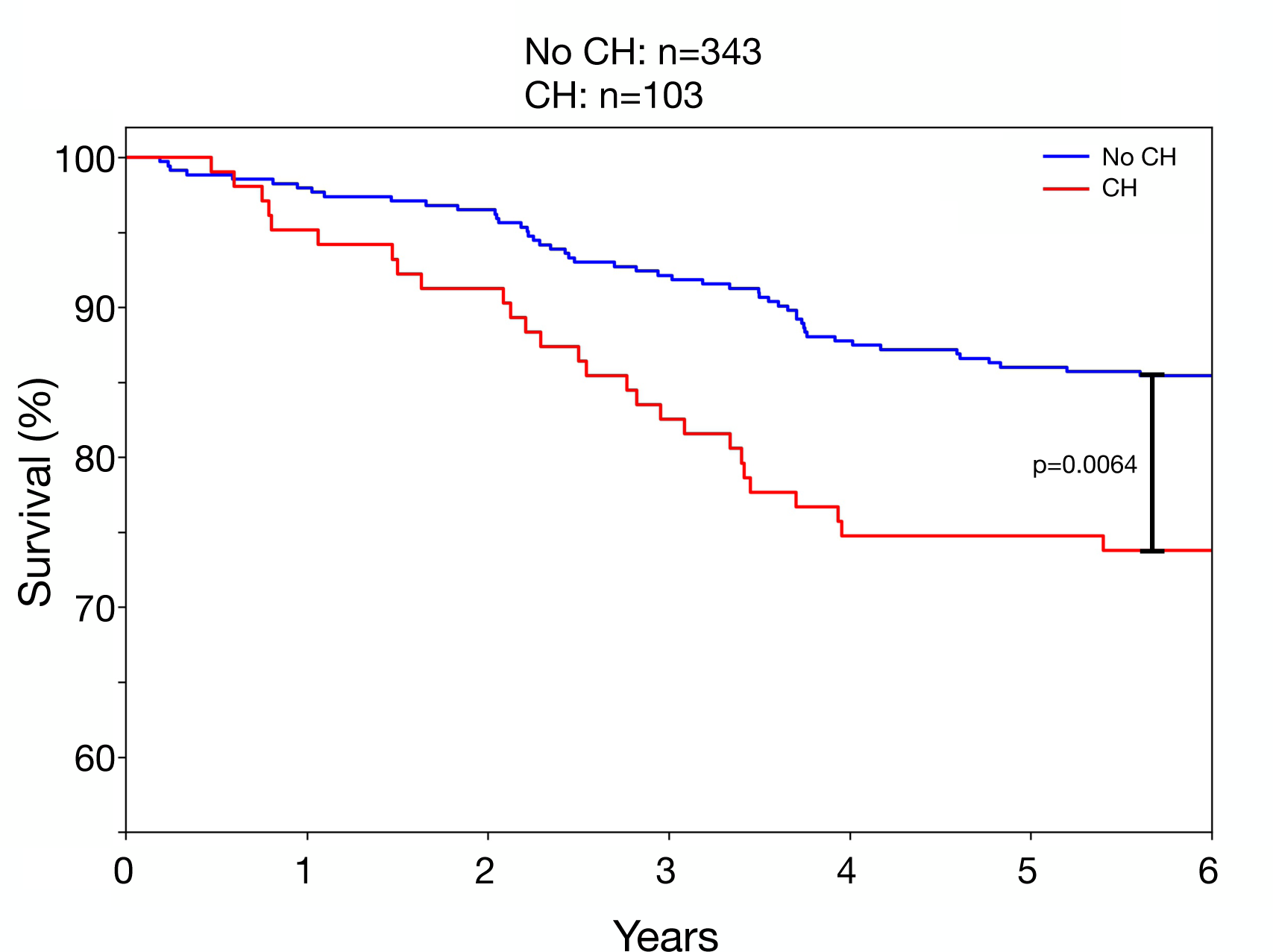
CH is associated with increased risk for mortality in patients with HFrEF. Kaplan-Meier survival curves showing association of CH status with incidence of mortality in patients with HFrEF. Cut-off levels for variant allele frequency was 1,15% for DNMT3A mutations and 0,73% for TET2 mutations. P-value was determined with log-rank test.

**Table 2:**
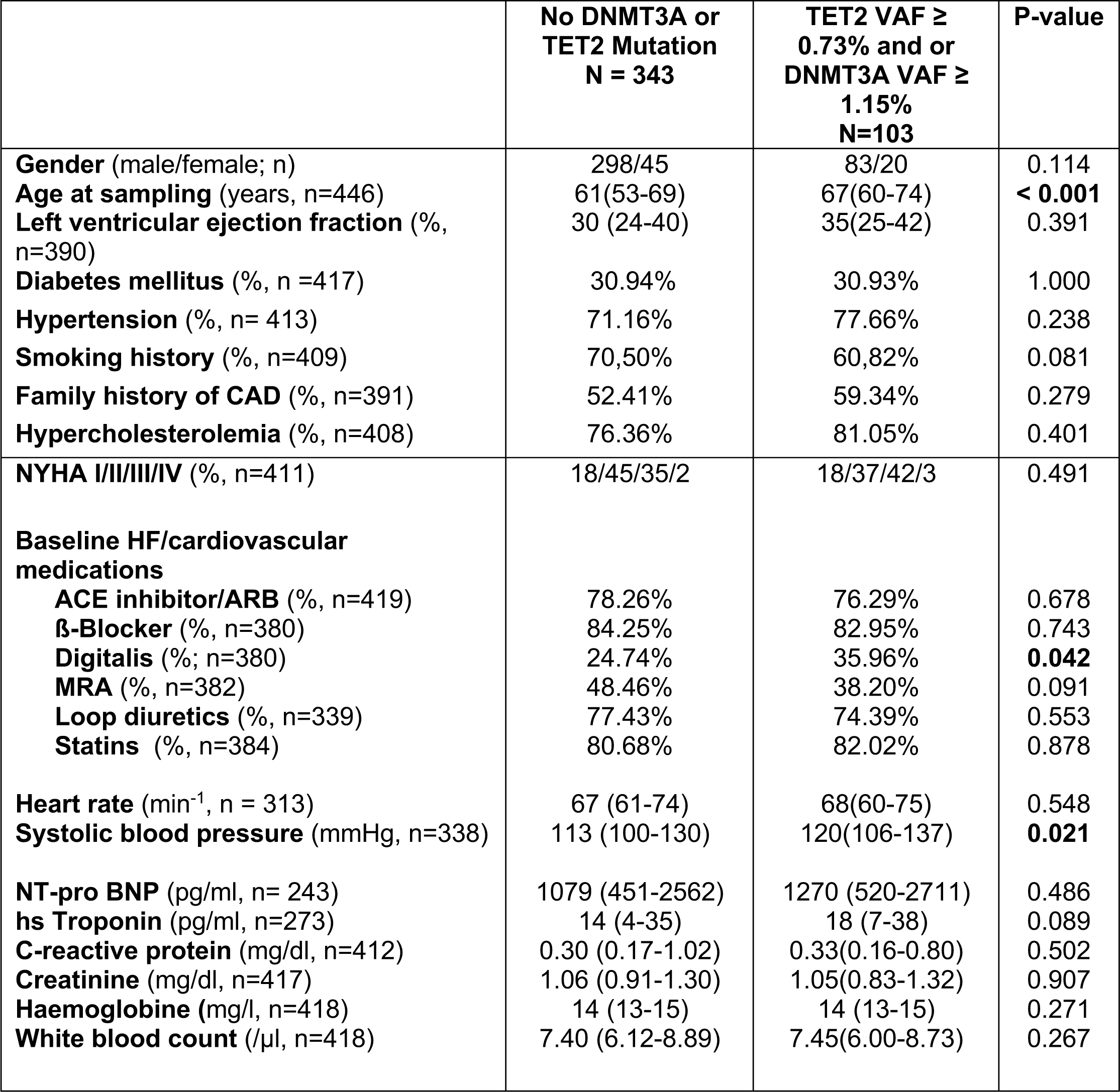
Baseline characteristics of the entire study cohort. Data are presented as median (interquartile range) for continuous variables. Categorical variables are shown as frequency (%). Significant values are shown in bold numbers.

| | No DNMT3A or<br>TET2 Mutation<br>N = 343 | TET2 VAF ≥<br>0.73% and or<br>DNMT3A VAF ≥<br>1.15%<br>N=103 | P-value |
| --- | --- | --- | --- |
| **Gender** (male/female; n) | 298/45 | 83/20 | 0.114 |
| **Age at sampling** (years, n=446) | 61(53-69) | 67(60-74) | **< 0.001** |
| **Left ventricular ejection fraction** (%<br>n=390) | 30 (24-40) | 35(25-42) | 0.391 |
| **Diabetes mellitus** (%<br>n =417) | 30.94% | 30.93% | 1.000 |
| **Hypertension** (%<br>n= 413) | 71.16% | 77.66% | 0.238 |
| **Smoking history** (%<br>n=409) | 70,50% | 60,82% | 0.081 |
| **Family history of CAD** (%<br>n=391) | 52.41% | 59.34% | 0.279 |
| **Hypercholesterolemia** (%<br>n=408) | 76.36% | 81.05% | 0.401 |
| **NYHA I/II/III/IV** (%<br>n=411) | 18/45/35/2 | 18/37/42/3 | 0.491 |
| **Baseline HF/cardiovascular**<br>**medications** |  |  |  |
| **ACE inhibitor/ARB** (%<br>n=419) | 78.26% | 76.29% | 0.678 |
| **β-Blocker** (%<br>n=380) | 84.25% | 82.95% | 0.743 |
| **Digitalis** (%<br>n=380) | 24.74% | 35.96% | **0.042** |
| **MRA** (%<br>n=382) | 48.46% | 38.20% | 0.091 |
| **Loop diuretics** (%<br>n=339) | 77.43% | 74.39% | 0.553 |
| **Statins** (%<br>n=384) | 80.68% | 82.02% | 0.878 |
| **Heart rate** (min<sup>-1</sup><br>n = 313) | 67 (61-74) | 68(60-75) | 0.548 |
| **Systolic blood pressure** (mmHg,<br>n=338) | 113 (100-130) | 120(106-137) | **0.021** |
| **NT-pro BNP** (pg/ml,<br>n= 243) | 1079 (451-2562) | 1270 (520-2711) | 0.486 |
| **hs Troponin** (pg/ml,<br>n=273) | 14 (4-35) | 18 (7-38) | 0.089 |
| **C-reactive protein** (mg/dl,<br>n=412) | 0.30 (0.17-1.02) | 0.33(0.16-0.80) | 0.502 |
| **Creatinine** (mg/dl,<br>n=417) | 1.06 (0.91-1.30) | 1.05(0.83-1.32) | 0.907 |
| **Haemoglobine** (mg/l,<br>n=418) | 14 (13-15) | 14 (13-15) | 0.271 |
| **White blood count** (/μl,<br>n=418) | 7.40 (6.12-8.89) | 7.45(6.00-8.73) | 0.267 |

### Association of SNPs with abrogation of increased mortality risk in CH carriers

Out of the preselected SNPs **(Table1),** we identified three common genetic variants, which were associated with abrogation of the increased risk for mortality in carriers of DNMT3A or TET2 driver mutations. All three variants were located in the IL6-receptor gene, were in strong linkage disequilibrium with each other and occurred in 62-65% of patients carrying a CH-driver mutation. As illustrated by Kaplan-Meier analysis in **Figure 2A-C**, rs2228145, rs4129267 and rs4537545 significantly attenuated the increased risk for mortality in carriers of CH-driver mutations, but had no effect in patients without DNMT3A or TET2 CH-driver mutations. Importantly, there were no significant differences in baseline clinical characteristics and pharmacological HFrEF treatment between patients with CH harboring the respective genetic variants compared to those without the respective SNPs (**Tables 3-5**), except for a slightly lower incidence of diabetes for the rs2228145 and rs4129267 variant. Moreover, the prevalence of all three SNPs did not differ between carriers of the CH-driver mutations and patients without CH-driver mutations in the DNMT3A or TET2 gene. Since all three SNPs, which mostly cooccurred in the same patients (**Figure 2D**), are regarded to serve as a proxy for IL-6 inhibition, these data suggest that inherited genetic variants disrupting IL-6 signaling abrogate the increased risk for fatal outcome in patients with HFrEF carrying CH driver mutations.

**Figure 2:**
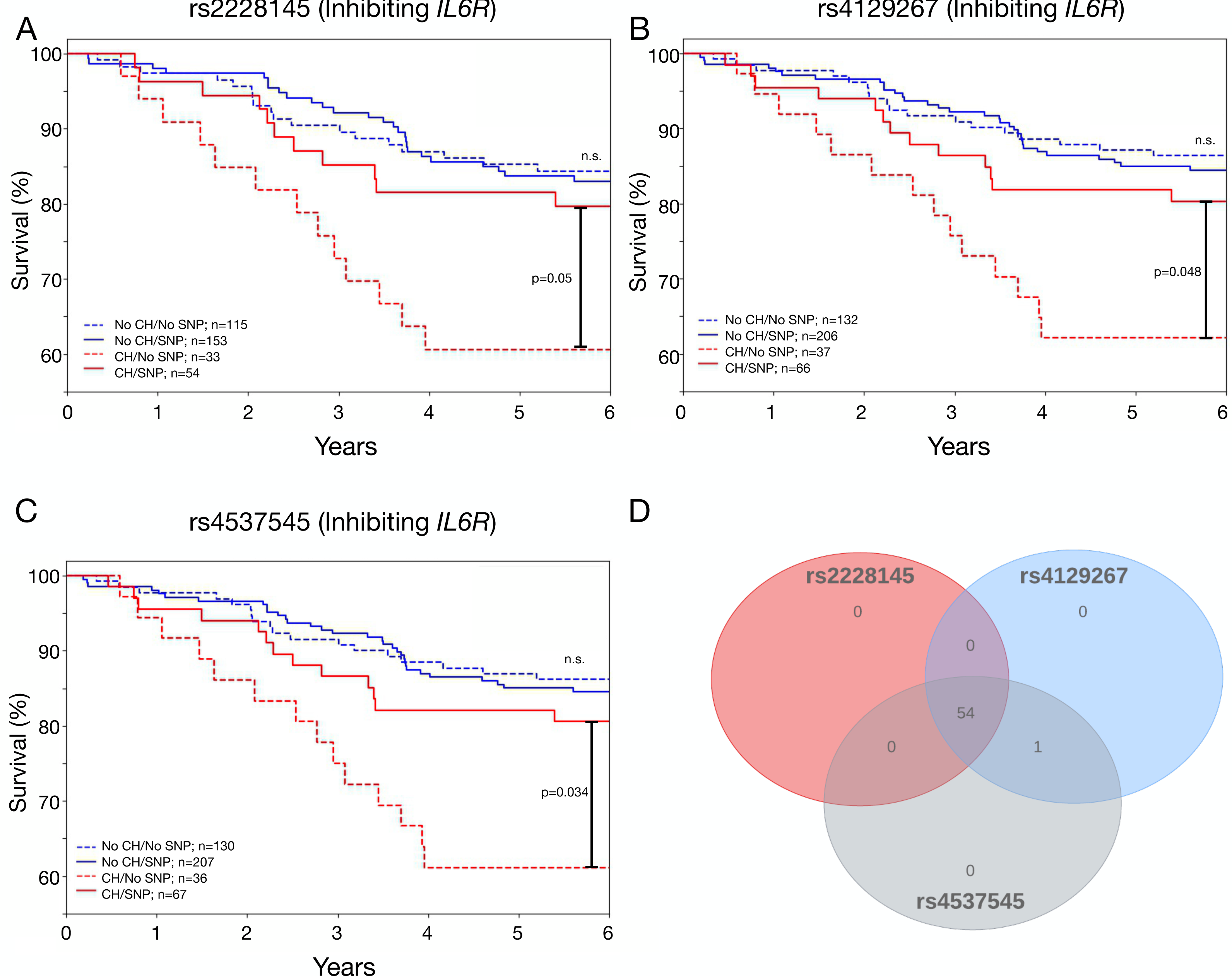
rs2228145, rs4129267 and rs453745 are associated with attenuation of risk for death in patients with HFrEF and CH. **A-C:** Kaplan Meier analysis showing association of CH status with or without the presence of rs2228145, rs4129267 and rs453745 with incidence of death in patients with HFrEF. N-numbers of included patients above the 90% confidence threshold for variant calling from the SNP array results are provided in each graph. P-values were determined with log-rank test. **D:** Venn diagram of patients with CH and the three prognosis improving SNPs demonstrating strong overlap of the three variants in patients with CH and HFrEF. Included were only patients, in whom all three variants were called.

**Table 3:**
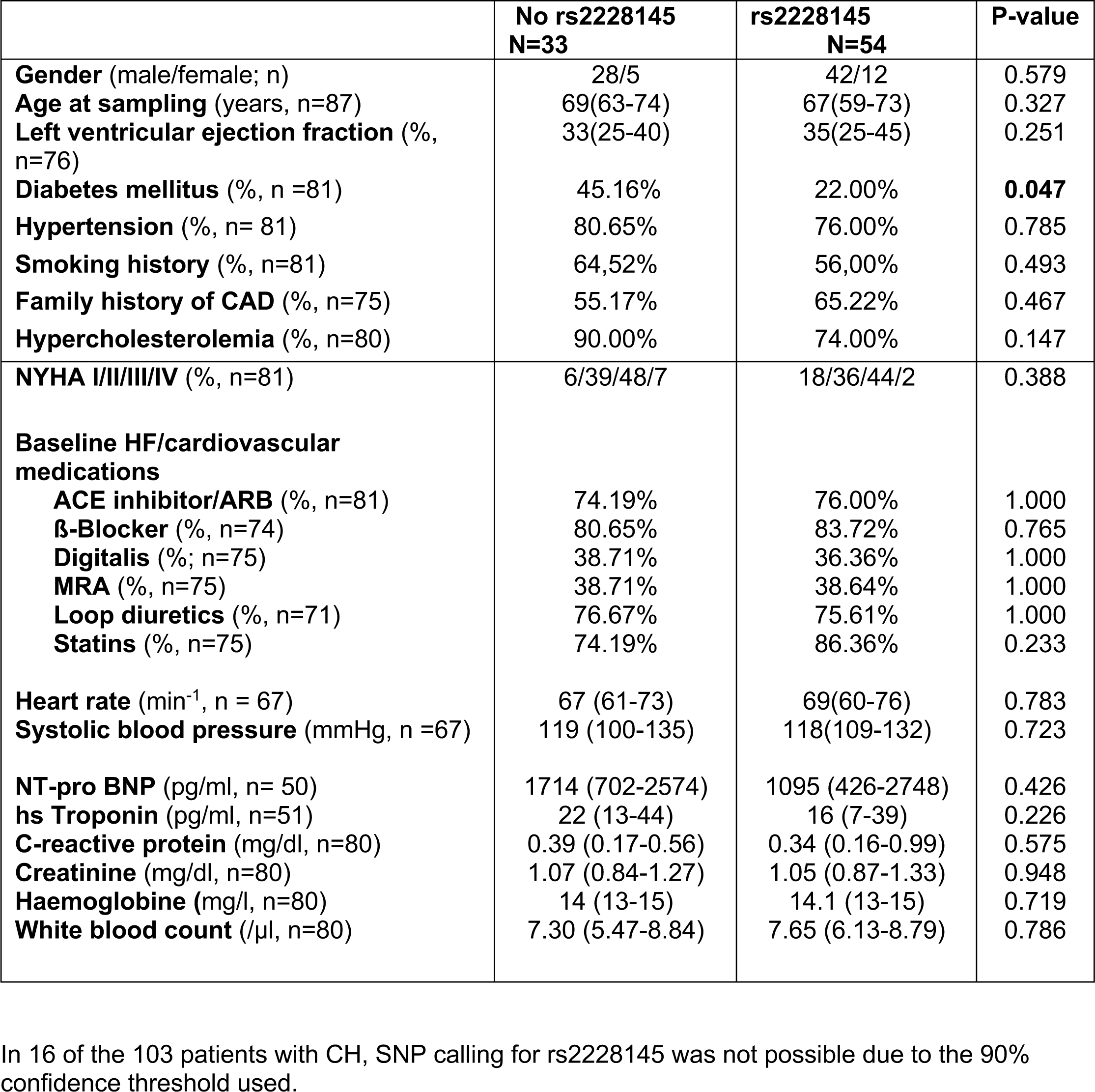
Baseline characteristics of patients with CH with or without the presence of rs2228145. Data are presented as median (interquartile range) for continuous variables. Categorical variables are shown as frequency (%). Significant values are shown in bold numbers.

|  | **No rs2228145**<br>**N=33** | **rs2228145**<br>**N=54** | **P-value** |
| --- | --- | --- | --- |
| **Gender** (male/female; n) | 28/5 | 42/12 | 0.579 |
| **Age at sampling** (years, n=87) | 69(63-74) | 67(59-73) | 0.327 |
| **Left ventricular ejection fraction** (%<br>n=76) | 33(25-40) | 35(25-45) | 0.251 |
| **Diabetes mellitus** (%<br>n =81) | 45.16% | 22.00% | **0.047** |
| **Hypertension** (%<br>n= 81) | 80.65% | 76.00% | 0.785 |
| **Smoking history** (%<br>n=81) | 64,52% | 56,00% | 0.493 |
| **Family history of CAD** (%<br>n=75) | 55.17% | 65.22% | 0.467 |
| **Hypercholesterolemia** (%<br>n=80) | 90.00% | 74.00% | 0.147 |
| **NYHA I/II/III/IV** (%<br>n=81) | 6/39/48/7 | 18/36/44/2 | 0.388 |
| **Baseline HF/cardiovascular**<br>**medications** |  |  |  |
| **ACE inhibitor/ARB** (%<br>n=81) | 74.19% | 76.00% | 1.000 |
| **β-Blocker** (%<br>n=74) | 80.65% | 83.72% | 0.765 |
| **Digitalis** (%<br>n=75) | 38.71% | 36.36% | 1.000 |
| **MRA** (%<br>n=75) | 38.71% | 38.64% | 1.000 |
| **Loop diuretics** (%<br>n=71) | 76.67% | 75.61% | 1.000 |
| **Statins** (%<br>n=75) | 74.19% | 86.36% | 0.233 |
| **Heart rate** (min<sup>-1</sup><br>n = 67) | 67 (61-73) | 69(60-76) | 0.783 |
| **Systolic blood pressure** (mmHg,<br>n =67) | 119 (100-135) | 118(109-132) | 0.723 |
| **NT-pro BNP** (pg/ml,<br>n= 50) | 1714 (702-2574) | 1095 (426-2748) | 0.426 |
| **hs Troponin** (pg/ml,<br>n=51) | 22 (13-44) | 16 (7-39) | 0.226 |
| **C-reactive protein** (mg/dl,<br>n=80) | 0.39 (0.17-0.56) | 0.34 (0.16-0.99) | 0.575 |
| **Creatinine** (mg/dl,<br>n=80) | 1.07 (0.84-1.27) | 1.05 (0.87-1.33) | 0.948 |
| **Haemoglobine** (mg/l,<br>n=80) | 14 (13-15) | 14.1 (13-15) | 0.719 |
| **White blood count** (/μl,<br>n=80) | 7.30 (5.47-8.84) | 7.65 (6.13-8.79) | 0.786 |

In 16 of the 103 patients with CH, SNP calling for rs2228145 was not possible due to the 90% confidence threshold used.

**Table 4:**
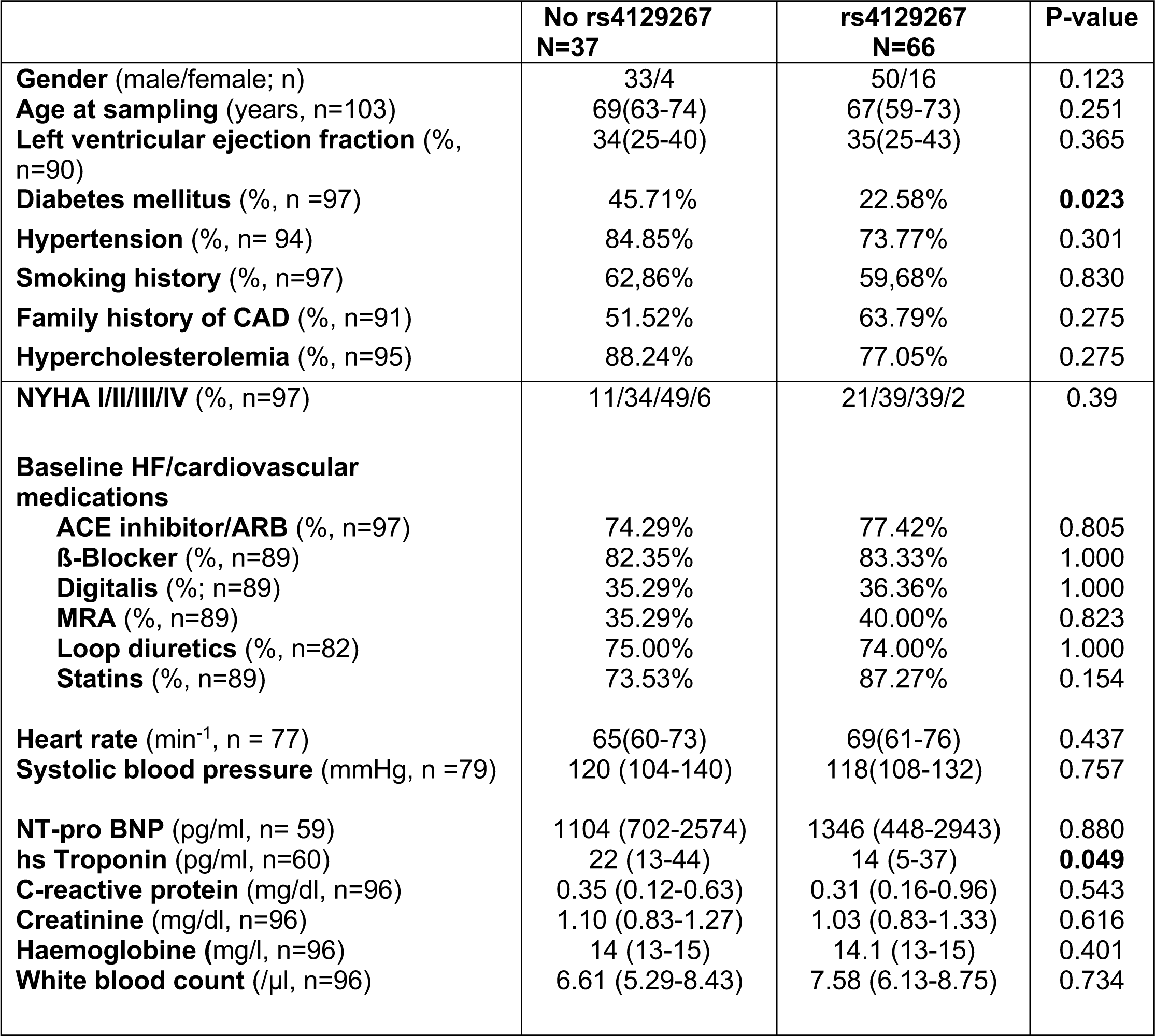
Baseline characteristics of patients with CH with or without the presence of rs4129267. Data are presented as median (interquartile range) for continuous variables. Categorical variables are shown as frequency (%). Significant values are shown in bold numbers.

|  | **No rs4129267**<br>**N=37** | **rs4129267**<br>**N=66** | **P-value** |
| --- | --- | --- | --- |
| **Gender** (male/female; n) | 33/4 | 50/16 | 0.123 |
| **Age at sampling** (years, n=103) | 69(63-74) | 67(59-73) | 0.251 |
| **Left ventricular ejection fraction** (%<br>n=90) | 34(25-40) | 35(25-43) | 0.365 |
| **Diabetes mellitus** (%<br>n =97) | 45.71% | 22.58% | **0.023** |
| **Hypertension** (%<br>n= 94) | 84.85% | 73.77% | 0.301 |
| **Smoking history** (%<br>n=97) | 62,86% | 59,68% | 0.830 |
| **Family history of CAD** (%<br>n=91) | 51.52% | 63.79% | 0.275 |
| **Hypercholesterolemia** (%<br>n=95) | 88.24% | 77.05% | 0.275 |
| **NYHA I/II/III/IV** (%<br>n=97) | 11/34/49/6 | 21/39/39/2 | 0.39 |
| **Baseline HF/cardiovascular**<br>**medications** |  |  |  |
| **ACE inhibitor/ARB** (%<br>n=97) | 74.29% | 77.42% | 0.805 |
| **β-Blocker** (%<br>n=89) | 82.35% | 83.33% | 1.000 |
| **Digitalis** (%<br>n=89) | 35.29% | 36.36% | 1.000 |
| **MRA** (%<br>n=89) | 35.29% | 40.00% | 0.823 |
| **Loop diuretics** (%<br>n=82) | 75.00% | 74.00% | 1.000 |
| **Statins** (%<br>n=89) | 73.53% | 87.27% | 0.154 |
| **Heart rate** (min<sup>-1</sup><br>n = 77) | 65(60-73) | 69(61-76) | 0.437 |
| **Systolic blood pressure** (mmHg,<br>n =79) | 120 (104-140) | 118(108-132) | 0.757 |
| **NT-pro BNP** (pg/ml,<br>n= 59) | 1104 (702-2574) | 1346 (448-2943) | 0.880 |
| **hs Troponin** (pg/ml,<br>n=60) | 22 (13-44) | 14 (5-37) | **0.049** |
| **C-reactive protein** (mg/dl,<br>n=96) | 0.35 (0.12-0.63) | 0.31 (0.16-0.96) | 0.543 |
| **Creatinine** (mg/dl,<br>n=96) | 1.10 (0.83-1.27) | 1.03 (0.83-1.33) | 0.616 |
| **Haemoglobine** (mg/l,<br>n=96) | 14 (13-15) | 14.1 (13-15) | 0.401 |
| **White blood count** (/μl,<br>n=96) | 6.61 (5.29-8.43) | 7.58 (6.13-8.75) | 0.734 |

**Table 5:** Baseline characteristics of patients with CH with or without the presence of rs4537545. Data are presented as median (interquartile range) for continuous variables. Categorical variables are shown as frequency (%). Significant values are shown in bold numbers.

|  | **No rs4537545**<br>**N=36** | **rs4537545**<br>**N=67** | **P-value** |
| --- | --- | --- | --- |
| **Gender** (male/female; n) | 32/4 | 51/16 | 0.190 |
| **Age at sampling** (years, n=103) | 69(63-74) | 67(59-73) | 0.177 |
| **Left ventricular ejection fraction** (%<br>n=90) | 34(25-40) | 35(25-43) | 0.377 |
| **Diabetes mellitus** (%<br>n =97) | 44.12% | 23.81% | 0.064 |
| **Hypertension** (%<br>n= 94) | 84.85% | 73.77% | 0.301 |
| **Smoking history** (%<br>n=97) | 64.71% | 58.73% | 0.665 |
| **Family history of CAD** (%<br>n=91) | 53.13% | 62.71% | 0.383 |
| **Hypercholesterolemia** (%<br>n=95) | 87.88% | 77.42% | 0.278 |
| **NYHA I/II/III/IV** (%<br>n=97) | 12/35/47/6 | 21/38/40/2 | 0.451 |
| **Baseline HF/cardiovascular**<br>**medications** |  |  |  |
| **ACE inhibitor/ARB** (%<br>n=97) | 73.53% | 77.78% | 0.628 |
| **β-Blocker** (%<br>n=88) | 81.82% | 83.64% | 1.000 |
| **Digitalis** (%<br>n=89) | 33.33% | 37.50% | 0.820 |
| **MRA** (%<br>n=89) | 36.36% | 39.29% | 0.825 |
| **Loop diuretics** (%<br>n=82) | 77.42% | 72.55% | 0.795 |
| **Statins** (%<br>n=89) | 72.73% | 87.27% | 0.154 |
| **Heart rate** (min<sup>-1</sup><br>n = 77) | 66(60-73) | 69(61-76) | 0.566 |
| **Systolic blood pressure** (mmHg,<br>n =79) | 120 (103-140) | 119(109-133) | 0.970 |
| **NT-pro BNP** (pg/ml,<br>n= 59) | 1092 (654-2570) | 1422(464-2865) | 1.000 |
| **hs Troponin** (pg/ml,<br>n=60) | 22 (13-44) | 15 (6-36) | 0.061 |
| **C-reactive protein** (mg/dl,<br>n=96) | 0.34 (0.11-0.67) | 0.31 (0.17-0.91) | 0.446 |
| **Creatinine** (mg/dl,<br>n=96) | 1.09 (0.82-1.27) | 1.04 (0.83-1.33) | 0.857 |
| **Haemoglobine** (mg/l,<br>n=96) | 14.00 (13-15) | 14.10 (13-15) | 0.319 |
| **White blood count** (/μl,<br>n=96) | 6.96 (5.22-8.64) | 7.53 (6.13-8.73) | 0.568 |

### Association of SNPs with increased mortality risk in CH carriers

On the contrary, we identified another three common genetic variants, which were associated with a significantly increased risk for fatal outcome in patients with HFrEF carrying CH driver mutations. As illustrated by Kaplan-Meier curves in **Figure 3A-C**, rs2250417, located in the beta-carotene oxygenase 2 gene close to the IL-18 gene (**Figure 3A**), rs4722172, an intergenic variant in the IL-6 gene (**Figure 3B**), and rs4845625, an intron variant in the IL-6 receptor gene (**Figure 3C**), all were associated with a significantly increased mortality in carriers of CH-driver mutations. Importantly, these three SNPs are not in strong linkage disequilibrium (**Figure 3D**). Their prevalence in carriers of CH-driver mutations varied from 69% for the rs2250417 and rs4845625 variants to 30% for the rs4722172 variant. The baseline clinical characteristics and pharmacological therapy was similar for CH driver mutation carriers with or without harboring the respective variants, except for a slightly lower systolic blood pressure with the rs2250417 variant and a slightly higher incidence of NYHA II stage heart failure in the rs4845625 SNP variant group (**Tables 6-8**). None of the variants was associated with altered mortality in HFrEF patients without CH driver mutations. As the rs2250417 variant associates with increased IL-18 levels, an important upstream mediator in inflammasome mediated innate immune responses, and rs4722172 and rs4845625 associate with enhanced IL-6 signaling, these data suggest that an inherited genetic susceptibility to inflammatory pathway activation may pave the way for an exuberant proinflammatory activation during acquisition of CH-driver mutations, accounting in large part for the increased mortality in HFrEF with CH.

**Figure 3:**
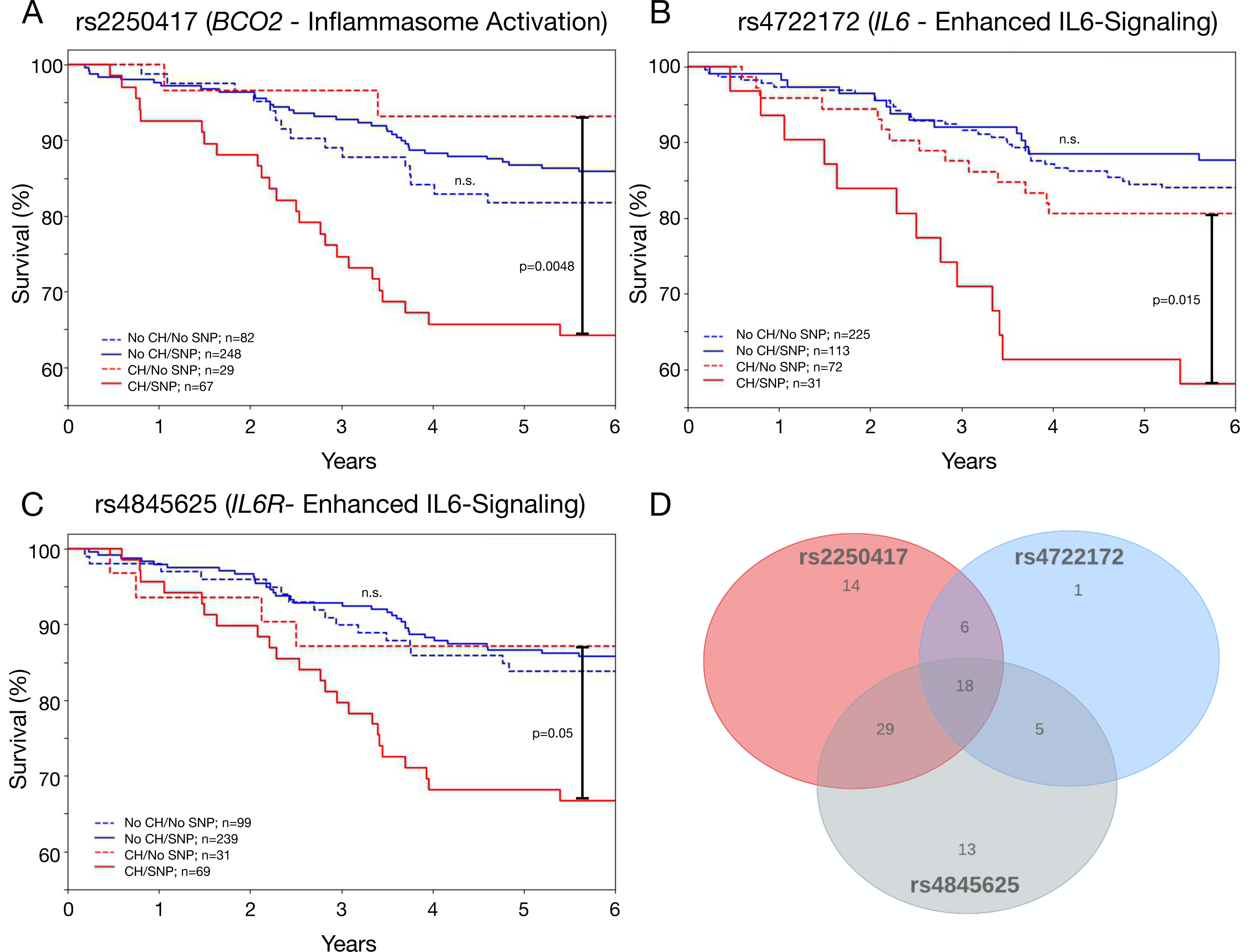
rs2250417, rs4722172 and rs4845625 are associated with increased mortality risk in patients with HFrEF and CH. **A-C:** Kaplan Meier analysis showing association of CH status with or without the presence of rs2250417, rs4722172 and rs4845625 with incidence of death in patients with HFrEF. N-numbers of included patients above the 90% confidence threshold for variant calling from the SNP array results are provided in each graph. P-values were determined with log-rank test. **D:** Venn diagram of patients with CH and the three prognosis aggravating SNPs demonstrating only weak overlap of the three variants in patients with CH and HFrEF. Included were only patients in which all three variants were called.

**Table 6:** Baseline characteristics of patients with CH with or without the presence of rs2250417. Data are presented as median (interquartile range) for continuous variables. Categorical variables are shown as frequency (%). Significant values are shown in bold numbers.

|  | **No rs2250417**<br>**N=29** | **rs2250417**<br>**N=67** | **P-value** |
| --- | --- | --- | --- |
| **Gender** (male/female; n) | 23/6 | 54/13 | 1.000 |
| **Age at sampling** (years, n=96) | 71(60-76) | 66(60-73) | 0.119 |
| **Left ventricular ejection fraction** (%<br>n=85) | 39(34-45) | 31(25-42) | 0.056 |
| **Diabetes mellitus** (%<br>n =91) | 40.74% | 26.56% | 0.217 |
| **Hypertension** (%<br>n= 90) | 80.77% | 76.56% | 0.784 |
| **Smoking history** (%<br>n=91) | 70.37% | 56.25% | 0.246 |
| **Family history of CAD** (%<br>n=86) | 60.00% | 62.30% | 1.000 |
| **Hypercholesterolemia** (%<br>n=89) | 88.46% | 79.37% | 0.378 |
| **NYHA I/II/III/IV** (%<br>n=91) | 19/33/44/4 | 14/39/44/3 | 0.933 |
| **Baseline HF/cardiovascular**<br>**medications** |  |  |  |
| **ACE inhibitor/ARB** (%<br>n=91) | 70.37% | 79.69% | 0.415 |
| **β-Blocker** (%<br>n=83) | 92.00% | 79.31% | 0.210 |
| **Digitalis** (%<br>n=84) | 30.77% | 41.38% | 0.820 |
| **MRA** (%<br>n=84) | 23.08% | 46.55% | 0.054 |
| **Loop diuretics** (%<br>n=80) | 68.00% | 76.36% | 0.428 |
| **Statins** (%<br>n=89) | 76.92% | 84.48% | 0.538 |
| **Heart rate** (min<sup>-1</sup><br>n = 75) | 63(59-72) | 69(61-75) | 0.281 |
| **Systolic blood pressure** (mmHg,<br>n =77) | 131 (112-140) | 114(105-130) | **0.029** |
| **NT-pro BNP** (pg/ml,<br>n= 55) | 997 (433-2574) | 1459(574-2831) | 0.332 |
| **hs Troponin** (pg/ml,<br>n=58) | 18 (7-35) | 18 (8-34) | 0.911 |
| **C-reactive protein** (mg/dl,<br>n=90) | 0.35 (0.17-0.63) | 0.33 (0.14-0.80) | 0.959 |
| **Creatinine** (mg/dl,<br>n=90) | 1.02 (0.8-1.33) | 1.10 (0.89-1.33) | 0.365 |
| **Haemoglobine** (mg/l,<br>n=90) | 14.0(13.9-15) | 14.00 (13.00-14.95) | 0.139 |
| **White blood count** (/μl,<br>n=90) | 7.26 (6.0-8.1) | 7.49 (6.0-8.91) | 0.427 |

In 7 of the 103 patients with CH, SNP calling for rs2250417 was not possible due to the 90% confidence threshold used.

**Table 7:** Baseline characteristics of patients with CH with or without the presence of rs4722172. Data are presented as median (interquartile range) for continuous variables. Categorical variables are shown as frequency (%). Significant values are shown in bold numbers.

|  | **No rs4722172**<br>**N=72** | **rs4722172**<br>**N=31** | **P-value** |
| --- | --- | --- | --- |
| **Gender** (male/female; n) | 59/13 | 24/7 | 0.596 |
| **Age at sampling** (years, n=103) | 68(60-75) | 66(60-73) | 0.119 |
| **Left ventricular ejection fraction** (%<br>n=90) | 39(34-45) | 30(23-43) | 0.327 |
| **Diabetes mellitus** (%<br>n =97) | 28.36% | 36.67% | 0.479 |
| **Hypertension** (%<br>n = 94) | 78.13% | 76.67% | 1.000 |
| **Smoking history** (%<br>n=97) | 59.70% | 63.33% | 0.824 |
| **Family history of CAD** (%<br>n=91) | 53.23% | 72.41% | 0.110 |
| **Hypercholesterolemia** (%<br>n=95) | 83.33% | 75.86% | 0.406 |
| **NYHA I/II/III/IV** (%<br>n=97) | 19/34/42/5 | 14/43/43/0 | 0.523 |
| **Baseline HF/cardiovascular**<br>**medications** |  |  |  |
| **ACE inhibitor/ARB** (%<br>n=97) | 77.61% | 73.33% | 0.797 |
| **β-Blocker** (%<br>n=88) | 85.00% | 78.57% | 0.545 |
| **Digitalis** (%<br>n=89) | 37.70% | 32.14% | 0.643 |
| **MRA** (%<br>n=89) | 34.43% | 46.43% | 0.349 |
| **Loop diuretics** (%<br>n=82) | 80.70% | 60.00% | 0.059 |
| **Statins** (%<br>n=89) | 81.97% | 82.14% | 1.000 |
| **Heart rate** (min<sup>-1</sup><br>n = 77) | 68(60-73) | 69(61-79) | 0.276 |
| **Systolic blood pressure** (mmHg<br>n =79) | 118 (103-132) | 121(110-141) | 0.135 |
| **NT-pro BNP** (pg/ml<br>n= 59) | 1307 (495-2711) | 1270(568-2755) | 0.891 |
| **hs Troponin** (pg/ml<br>n=60) | 18 (8-43) | 18 (7-30) | 0.448 |
| **C-reactive protein** (mg/dl<br>n=96) | 0.32 (0.16-0.78) | 0.35 (0.18-0.88) | 0.711 |
| **Creatinine** (mg/dl<br>n=96) | 1.05 (0.81-1.33) | 1.11 (0.96-1.28) | 0.607 |
| **Haemoglobine** (mg/l<br>n=96) | 14 (13-15) | 14 (13.6-15) | 0.404 |
| **White blood count** (/μl<br>n=96) | 7.11 (6.0-8.34) | 7.63 (5.74-8.97) | 0.551 |

**Table 8:**
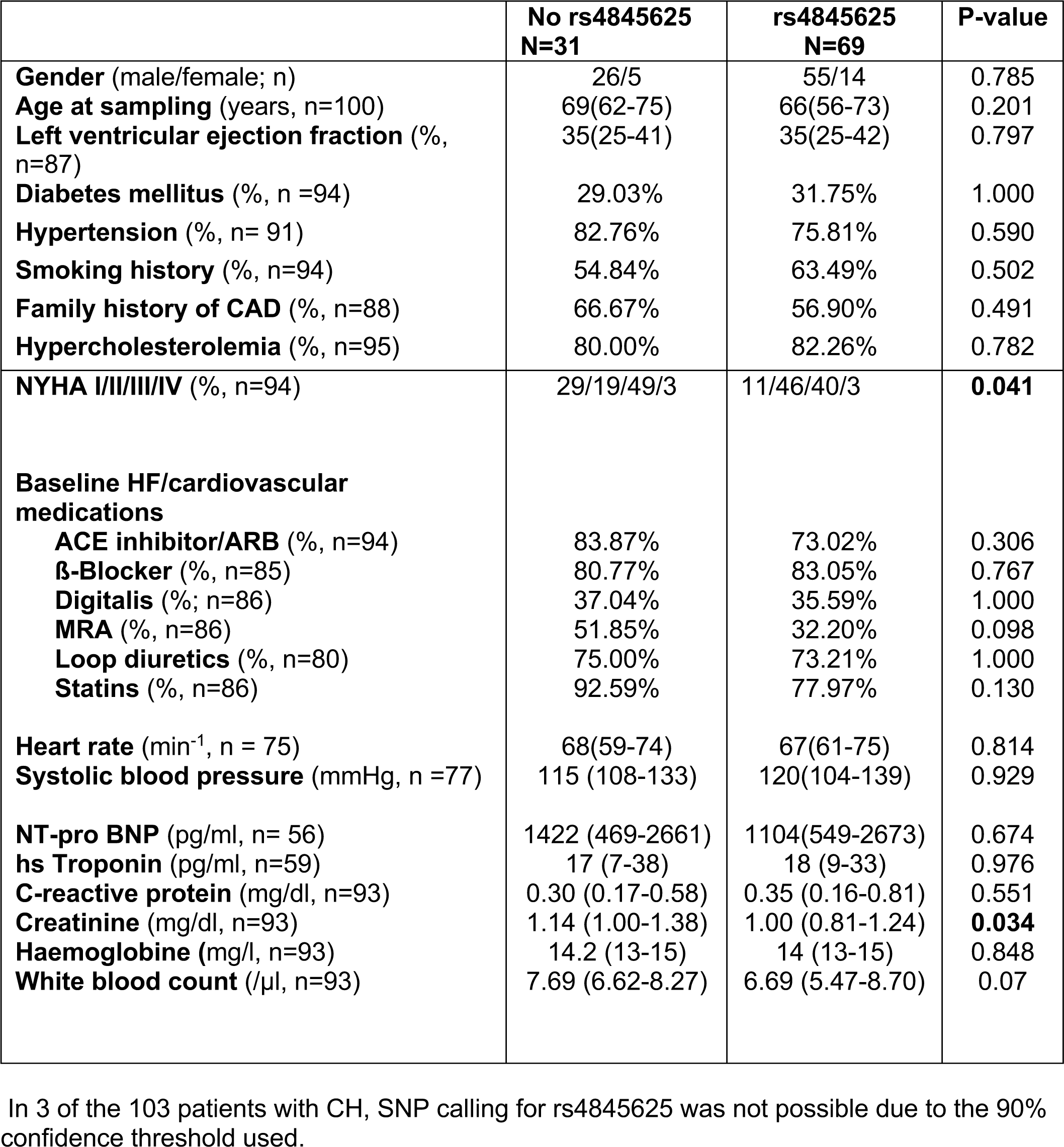
Baseline characteristics of patients with CH with or without the presence of rs4845625. Data are presented as median (interquartile range) for continuous variables. Categorical variables are shown as frequency (%). Significant values are shown in bold numbers.

### Proinflammatory gene signature in monocytes from DNMT3A-CH-mutation carriers harboring SNPs associated with increased mortality

In order to substantiate our findings and provide for a mechanistic underpinning for the hypothesis that an inherited genetic susceptibility drives proinflammatory activation in HFrEF patients carrying CH driver mutations, we performed single cell RNA sequencing of circulating monocytes obtained from DNMT3A mutation carriers harboring at least one mortality aggravating SNP, but no protecting SNPs compared to monocytes isolated from DNMT3A mutation carriers harboring SNPs, which abrogate increased mortality in CH-carriers (see **Table 9** for SNP class distribution). As illustrated in **Figure 4**, there was no difference in the distribution of classical and non-classical monocytes between the two groups. However, gene ontology pathway analysis revealed a profound and significant upregulation of proinflammatory genes in monocytes obtained from DNMT3A mutation carriers, who simultaneously harbored mortality aggravating genetic variants without having protective SNPs at the same time (**Figure 4E**). The top regulated genes were part of pathways of an enhanced innate immune response including cellular response to stress and cell activation as well as cytokine signaling. Thus, these data document that inherited genetic variants within the inflammasome-IL6 signaling pathway profoundly modulate the inflammatory gene signature of patients with HFrEF carrying DNMT3A CH-driver mutations.

**Figure 4:**
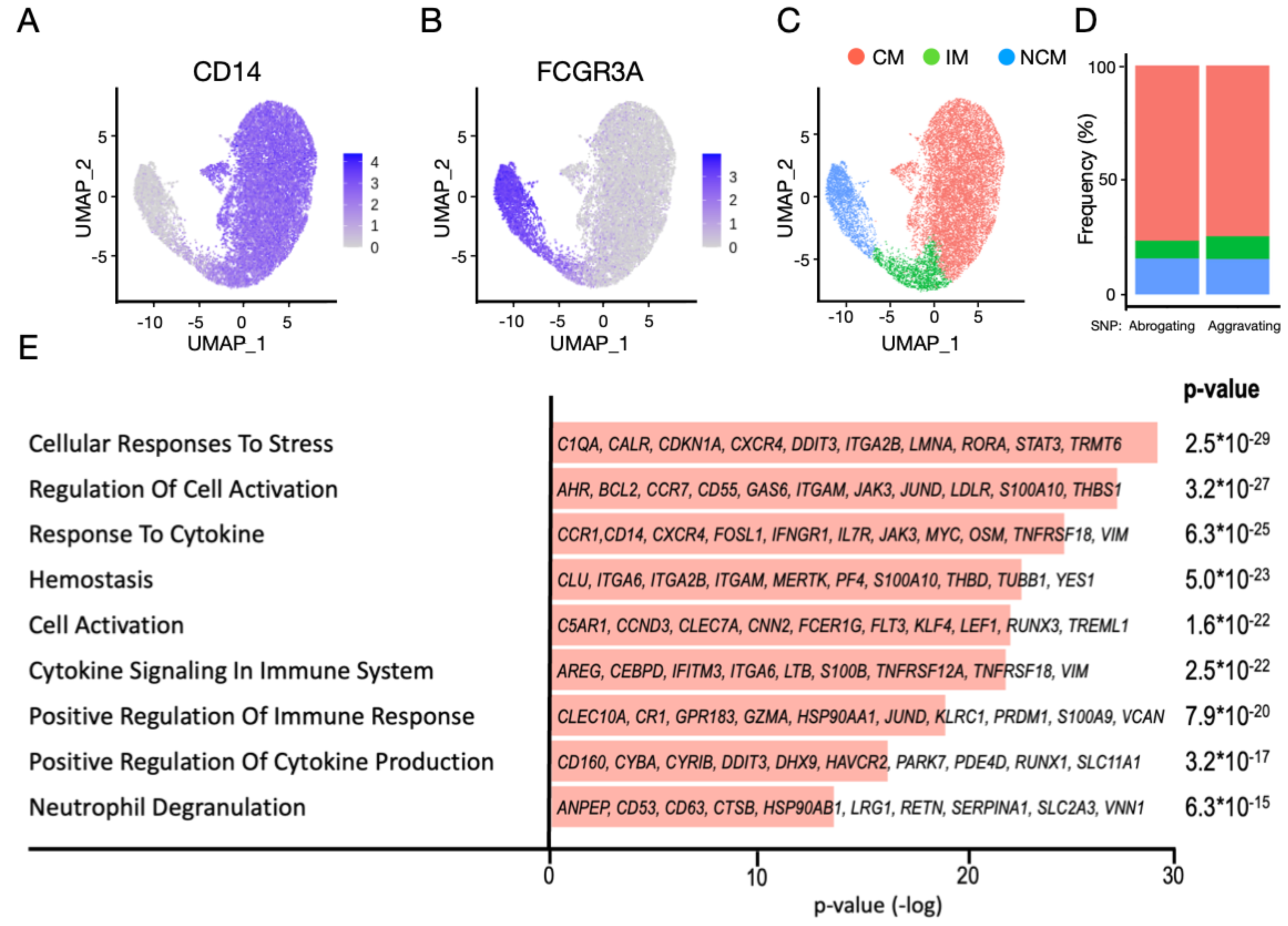
Single cell RNA-sequencing of circulating monocytes in patients with DNMT3A mutations according to SNP status. **A and B:** t-distributed Stochastic Neighbor Embedding (t-SNE) plot showing monocyte markers CD14 and FCGR3A (CD16). **C:** Distribution of classical, non-classical and intermediate monocytes in the samples analyzed. **D:** No difference in the proportion of monocyte subtypes in patients with DNMT3A mutations and aggravating or protecting SNPs. **E:** Pathway analysis showing upregulated pathways in patients with DNMT3A mutations and aggravating SNPs. Regulated genes in each pathway are highlighted. Significance was determined with a FDR test and p-values are provided. The reported terms are subset from summary terms that are reported from Metascape, with each summary term having similar nested terms of different source origins ( i.e. GO terms, KEGG pathways, Reactome). CM: Classcial Monocytes, IM: Intermediate Monocytes, NCM: Non-classical Monocytes.

**Table 9:** Distribution of SNPs in the single cell RNA-sequencing study. DNMT3A VAF describes variant allele frequency of the mutated CH clone. 1 denotes presence and 0 absence of a given SNP.

| **SNP Class** | **DNMT3A VAF** | **rs2228145** | **rs4129267** | **rs4537545** | **rs2250417** | **rs4722172** | **rs4845625** |
| --- | --- | --- | --- | --- | --- | --- | --- |
| Abrogating | 0,03748 | 1 | 1 | 1 | 0 | 0 | 1 |
| Abrogating | 0,04329 | 1 | 1 | 1 | No Call | 0 | 1 |
| Aggravating | 0,39447 | 0 | 0 | 0 | 0 | 0 | 1 |
| Aggravating | 0,05860 | 0 | 0 | 0 | 1 | 1 | 1 |
| Aggravating | 0,02407 | No Call | 0 | 0 | No Call | 0 | 1 |

## Discussion

The present study is the first to document that inherited genetic variants within the NLRP3 inflammasome to IL-6 signaling pathway profoundly modulate the increased risk for mortality in carriers of DNMT3A and TET2 driver mutations suffering from chronic HFrEF. Our data not only show that SNPs associated with disrupted IL-6 signaling abrogate the increased risk for fatal outcomes, but – more importantly – disclose that SNPs associated with enhanced inflammation in large part account for the increased mortality observed in carriers of DNMT3A and TET2 driver mutations. Mechanistically, our single cell RNA-sequencing data demonstrate that monocytes of patients with HFrEF carrying DNMT3A CH-driver mutations and simultaneously harboring IL-6 signaling enhancing genetic variants are characterized by a profound upregulation of genes involved in controlling immune and stress responses as well as cytokine activation. Thus, taken together, these data point towards a key role of IL-6 signaling to mediate the effects of CH on mortality in patients with HFrEF.

Our results showing that inherited genetic variants well established to be associated with disrupted IL-6 signaling modulate mortality risk in patients with HFrEF considerably extend the findings of a recent study showing that the rs2228145 IL-6 receptor variant^19^ abrogated the increased risk for the occurrence of atherosclerotic events in carriers of DNMT3A or TET2 CH-driver mutations.^17^ This observation has been reproduced in the complete 450 000-person UK biobank dataset using a stringent filtering strategy ^20, 21^. Importantly, the present study not only confirmed the association of the IL-6 receptor disrupting rs2228145 variant with an abrogated risk for fatal outcome, but also identified the rs4129267 IL-6 receptor variant to significantly reduce mortality in CH carriers with HFrEF. This variant is in strong linkage disequilibrium with the non-synonymous variant rs2228145. A very recent large Mendelian randomization study documented that the rs4129267 IL-6 receptor variant genetically determines elevated circulating soluble IL-6 receptor (sIL6R) plasma levels, which are causally linked with lower levels of hs-CRP, a sensitive biomarker of inflammation.^22^ Mechanistically, circulating sIL6R acts as a decoy receptor to neutralize IL-6. In addition, the third variant to abrogate the increased risk for mortality in patients with HFrEF carrying a CH driver mutation, rs4537545, located in the intron of the IL-6 receptor was recently shown to predict reduced circulating IL-6 levels as well as reduced hs-CRP levels.^23, 24^ Taken together, the results of the present study document that genetically determined reduced IL-6 signaling abrogates the increased mortality risk in patients with HFrEF carrying DNMT3A or TET2 CH driver mutations, thus providing robust evidence for the importance of proinflammatory circuits to mediate the increased risk for fatal outcome.

Likewise, and probably more important from a clinical point of view, we identified different inherited genetic variants, which are associated with an aggravated clinical outcome in patients with HFrEF carrying CH driver mutations. Two of these variants, rs47222172 as an intergenic variant in the IL-6 gene and rs4845625 as an intron variant of the IL-6 receptor, were previously identified as genetic risk loci for coronary artery disease^25, 26^ and peripheral artery disease,^27^ respectively. Of specific interest, the third variant we identified to aggravate fatal outcome in HFrEF patients carrying CH driver mutations, rs2250417, has so far not been implicated in cardiovascular disease and encodes for an intron variant in the beta-carotene oxygenase 2 gene close to the IL-18 gene.^28^ Importantly, the rs2250417 variant was shown to significantly associate with increased circulating IL-18 levels.^29^ Besides IL-1ß, IL-18 plays an essential role in the inflammasome mediated innate immune response ^30^ and elevated IL-18 plasma levels associate with increased cardiovascular death in patients with coronary artery disease.^31^

In support of a prominent role for the inherited genetic variants associated with activation of the NLRP3 inflammasome-IL-6 pathway to aggravate fatal outcome in HFrEF patients carrying CH driver mutations, our single cell RNA-sequencing data demonstrated that monocytes obtained from carriers of a DNMT3A CH-driver mutation, who simultaneously harbored at least one of the three prognosis aggravating SNPs without having protecting SNPs, displayed a proinflammatory gene signature involving all aspects of an enhanced innate immune response including the prototypical IL-6 activated transcription factor, STAT3. As such, the present study not only discloses a key role for the NLRP3 inflammasome-IL-6 pathway to modulate the increased risk for mortality in HFrEF patients carrying CH driver-mutations, but may also help to identify high-risk patients, who are specifically amenable to anti-inflammatory therapies.

Of note, the inherited genetic variants interfering with the NLRP3-IL-6 pathway investigated in the present study did not affect prognosis in the absence of CH driver mutations in our patient cohort with chronic HFrEF. Therefore, we hypothesize that the acquisition of somatic mutations during lifetime leading to CH sensitizes circulating immune cells to an exuberant proinflammatory activation in the presence of genetic variants associated with activation of the NLRP3-IL-6 pathway. Indeed, a previous study demonstrated that the risk for coronary heart disease as well as all-cause mortality in a selected population without previous atherosclerotic events was largely confined to CH mutation carriers, who additionally had evidence for accelerated epigenetic aging.^32^ Thus, it appears that CH requires as second hit to exert its detrimental effects on clinical outcome in cardiovascular diseases. The results of the present study indicate that an inherited genetic susceptibility to inflammatory pathway activation importantly contributes to the increased fatal outcome in patients with HFrEF carrying CH driver mutations. As such, the identification of inherited variants that modulate CH-related risk may be exquisitely suited to identify high-risk patients for targeted anti-inflammatory therapies. While a recent clinical trial employing antibodies directed against IL-1ß provided proof-of concept for anti-inflammatory therapies to reduce atherothrombotic complications, including hospitalization for heart failure in patients with established coronary artery disease,^33, 34^ treatments specifically directed at inflammation in patients with chronic HFrEF did not show improved clinical outcome.^15^ In addition, IL-1ß inhibition was associated with a small, but significant increase in fatal infections.^33^ Thus, more precise implementation of anti-inflammatory therapies to patients with evidence for increased risk due to inflammation appears to be mandatory. The results of the present study suggest that inherited variants in the IL-6 pathway may prove to be especially suitable to enrich for high-risk subgroups of patients carrying CH driver mutations. Importantly, even after successful IL-1ß inhibition with canakinumab, there remains substantial residual inflammatory risk, which was shown to be related to levels of IL18 and IL-6.^35^ Numerous anti IL-6 therapeutics are currently being tested in cardiovascular disease,^36^ which may be specifically useful for precision targeting inflammatory circuits in carriers of CH driver mutations harboring activating IL-6 signaling genetic variants, but may be futile in CH carriers with HFrEF and IL-6 signaling disrupting SNPs.

### Limitations

The present study is a retrospective analysis with a limited sample size from a single centre. However, survival data from the underlying clinical trials were already entered into locked databases when genetic analysis was performed. Also, the conclusions drawn from the single cell sequencing analysis are rather preliminary, as sample size is small. Therefore, these findings need validation in larger cohorts. Furthermore, due to the sensitivity of the Fluidigm assay, precise determination of SNP status was not possible for all inherited variants investigated. For example, a call for rs228145 was only possible in 84,4% of all patients with CH. However, we assume that our results are valid, as this SNP is in strong linkage disequilibrium with rs4129267 and rs4537545, which could be determined in all patients with CH in our cohort. Finally, we focused on SNPs in the NLRP3 inflammasome/IL-6 signaling pathway, which should not rule out that inherited variants in other pathways mediating cardiovascular damage could modulate outcomes in patients with HFrEF and CH.

## Data Availability

All data produced in the present study are available upon reasonable request to the authors

## Funding

We acknowledge funding from the Alfons und Gertrud Kassel-Stiftung as part of the center for data science and AI to NK, MHS and AMZ. This work was supported by the DZHK (German Centre for Cardiovascular Research), the German research Foundation (Exc 2026/1), the Dr. Robert Schwiete Foundation (to SD), the Willy Robert Pitzer Stiftung (to SC) and the Jose Carreras Leukämie-Stiftung (grant DJCLS 11 R/2020 to M.A.R.).

---

**Supplementary Table 1:** Sequences used for SNP genotyping.

**Supplementary Table 2:** Distribution of SNPs on two different measuring Chips.

## Notes

### Competing Interest Statement

The authors have declared no competing interest.

