## Supplementary figures and images for "Interaction of inherited genetic variants in the NLRP3 inflammasome/IL-6 pathway with acquired clonal hematopoiesis to modulate mortality risk in patients with HFrEF"

### Supplemental Table 1

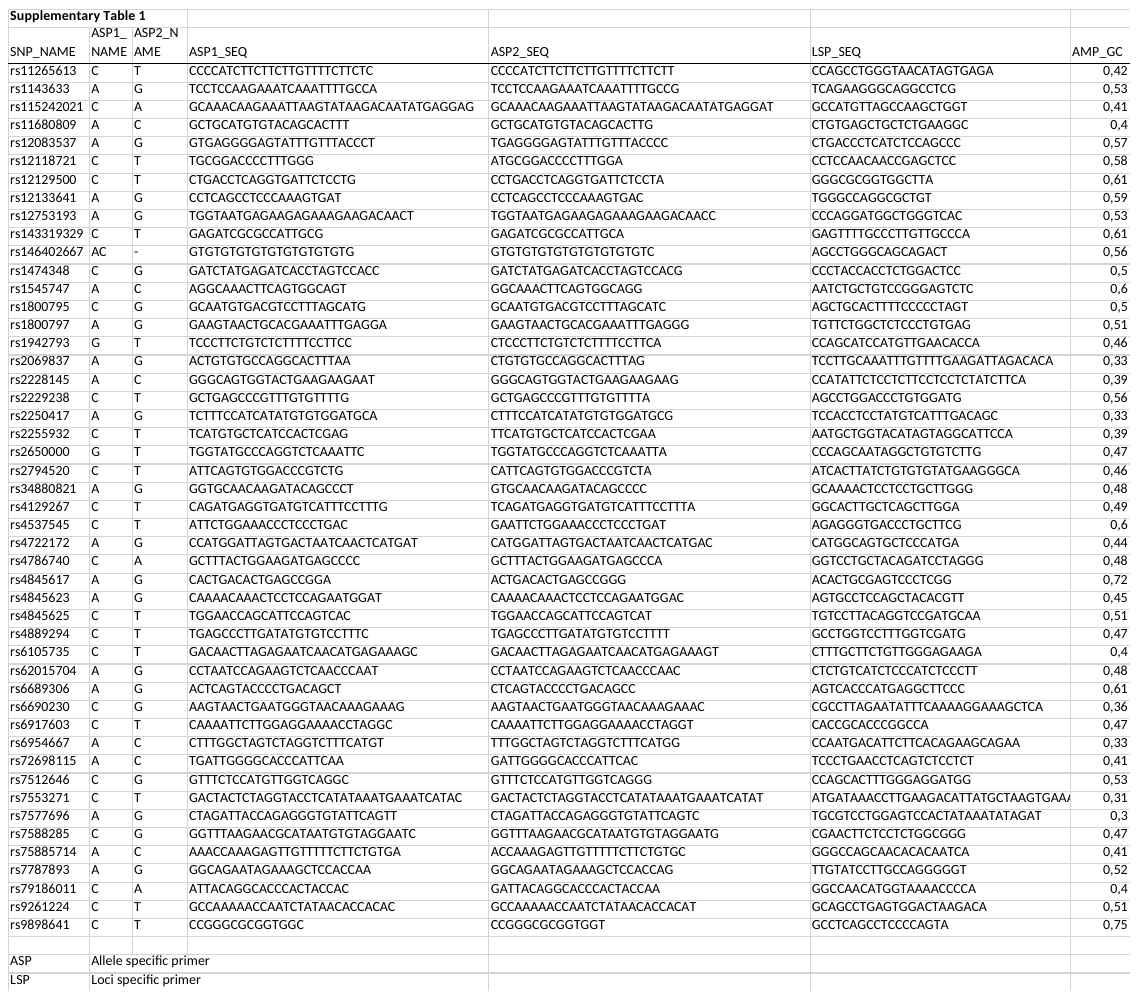
