## Supplemental Table 2 for "Interaction of inherited genetic variants in the NLRP3 inflammasome/IL-6 pathway with acquired clonal hematopoiesis to modulate mortality risk in patients with HFrEF"

| ***Supplementary Table 2*** | *Distribution of SNP assays on the IFC chips for measurements* | | |
| --- | --- | --- | --- |
| IFC1 | | IFC2 | |
| rs11265613 | IL6 | rs4129267 | IL6 |
| rs1143633 | IL1_CASP1 | rs4537545 | IL6 |
| rs115242021 | IL1_CASP1 | rs4722172 | IL6 |
| rs11680809 | IL1_CASP1 | rs4786740 | IL1_CASP1 |
| rs12083537 | IL6 | rs4845617 | IL6 |
| rs12118721 | IL6 | rs4845623 | IL6 |
| rs12129500 | IL6 | rs4845625 | IL6 |
| rs12133641 | IL6 | rs4889294 | IL1_CASP1 |
| rs12753193 | IL6 | rs6105735 | IL1_CASP1 |
| rs143319329 | IL1_CASP1 | rs62015704 | IL1_CASP1 |
| rs146402667 | IL6 | rs6689306 | IL6 |
| rs1474348 | IL6 | rs6690230 | IL6 |
| rs1545747 | IL1_CASP1 | rs6917603 | IL1_CASP1 |
| rs1800795 | IL6 | rs6954667 | IL6 |
| rs1800797 | IL6 | rs72698115 | IL6 |
| rs1942793 | IL1_CASP1 | rs7512646 | IL6 |
| rs2069837 | IL6 | rs7553271 | IL6 |
| rs2228145 | IL6 | rs7577696 | IL1_CASP1 |
| rs2229238 | IL6 | rs7588285 | IL1_CASP1 |
| rs2250417 | IL1_CASP1 | rs75885714 | IL1_CASP1 |
| rs2255932 | IL1_CASP1 | rs7787893 | IL6 |
| rs2650000 | IL6 | rs79186011 | IL1_CASP1 |
| rs2794520 | IL6 | rs9261224 | IL1_CASP1 |
| rs34880821 | IL6 | rs9898641 | IL1_CASP1 |
